# Negative Control Exposures: Causal effect Identifiability and Use in Probabilistic-Bias and Bayesian Analyses with Unmeasured Confounders

**DOI:** 10.1101/2022.05.25.22275304

**Authors:** WD Flanders, LA Waller, Q Zhang, D Getahun, M Silverberg, M Goodman

## Abstract

Probabilistic bias and Bayesian analyses are important tools for bias correction, particularly if required parameters are nonidentifiable. Negative controls are another tool; they can detect confounding and correct for confounders. Our goals are to present conditions that assure identifiability of certain causal effects and to describe and illustrate a probabilistic bias analysis and related Bayesian analysis that use a negative control exposure.

Using potential-outcome models, we characterize assumptions needed for identification of causal effects using a dichotomous, negative control exposure when residual confounding exists. We define bias parameters, characterize their relationships with the negative control and with specified causal effects, and describe the corresponding probabilistic-bias and Bayesian analyses.

We exemplify analyses using data on hormone therapy and suicide attempts among transgender people. To address possible confounding by healthcare utilization, we used prior TdaP (tetanus-diphtheria-pertussis) vaccination as a negative control exposure. Hormone therapy was weakly associated with risk (risk ratio (RR) = 0.9). The negative control exposure was associated with risk (RR = 1.7), suggesting confounding. Based on an assumed prior distribution for the bias parameter, the 95% simulation interval for the distribution of confounding-adjusted RR was (0.17, 1.64), with median 0.5; the 95% credibility interval was similar.

A dichotomous negative control exposure can be used to identify causal effects when a confounder is unmeasured under strong assumptions. More realistically, assumptions can be relaxed and the negative control exposure may prove helpful for probabilistic bias analyses and Bayesian analyses.

## Introduction

Residual confounding often threatens valid estimation of causal effects, especially absent randomization of exposure. In a potential outcome framework, confounding implies non-exchangeability, defined below as an association of the exposure with the potential outcomes.

Numerous approaches can adjust for, or otherwise account for measured confounders, including restriction to a single level of the confounder, control or adjustment by stratification or modelling in the analysis, difference in difference and regression discontinuity analyses, and use of instrumental variables^1^.

To detect residual confounding, perhaps due to an unmeasured or mis-measured confounder, one can use a *negative control outcome* or *negative control exposure*^2^. A negative control exposure, our focus, is a variable that does not cause the outcome but is associated with the suspected, but unmeasured confounder (detailed in methods). In an early application, Yerushalmy studied the effects of maternal cigarette smoking on birth weight (reprinted^3^). To detect confounding, he assessed the association of paternal smoking with his offspring’s birthweight; this alternative “exposure” was contemporaneously thought not to affect the outcome of interest – a negative control exposure; he observed an association and interpreted it as suggesting confounding. Later work using cotinine levels suggested this use of paternal smoking as a negative control was valid^2,4^. As another example, Flanders et al. studied the effects of air pollution on emergency department visits for respiratory diseases. They used pollutant levels the day after the outcome had occurred, which could not cause the outcome, as a negative control exposure to detect residual confounding or other bias^5,6^. A study of influenza vaccination and deaths from influenza^7^ exemplifies negative control outcomes. A strong association of vaccination status with influenza deaths *before* the influenza season (an alternative “outcome” thought to be unaffected by the exposure of interest – a negative control outcome) suggested bias in the estimated effect of vaccination. Lipsitch et al. discussed and formalized these concepts for detection of residual confounding^8^.

Much subsequent work goes beyond bias detection to use negative controls to adjust for residual confounding^2,9^. Flanders et al. used a negative control exposure to partially correct for residual confounding^10^. Their approach, however, involved certain distributional assumptions. Other approaches have involved outcome calibration under a rank preservation assumption^11^, and use of a linear model for the unmeasured confounder with factor analysis^12,13^. Miao et al. recently provided conditions that, if met, allow identification of causal effects by using two negative controls, which can act as surrogates for the unmeasured confounder(s)^14-16^.

Assumptions needed for identifiability can be rather strong if a confounder remains unmeasured. For example, in the categorical case, the approach of Miao et al.^14^ requires two negative controls that serve as proxies for the unmeasured confounder (say U). They must have several properties including: each proxy has at least as many categories as U, the proxies are independent conditional on U and certain probability matrices have inverses.

Probabilistic bias analyses can address residual biases in the effect estimate that remain after conventional analyses^17^. For residual confounding, probabilistic bias analyses use substantive knowledge to help formulate a distribution of bias parameters that characterize unobserved associations (specified in methods), apply that distribution to correct conventional effect estimates, such as risk ratios, and produce a distribution of plausible corrected estimates. Our goals are to describe and illustrate a method that uses a negative control exposure to partially correct for confounding and to formulate probabilistic bias analyses. We extend the approach to a fully Bayesian analysis.

## Methods

### Background, Notation, and Definitions

Our specific objectives are to: present and justify conditions sufficient for using a negative control exposure (*N*) to identify causal effects of an exposure (*E*) on an outcome (*Y*) when a confounder (*U*) is unmeasured; describe probabilistic bias analyses to address confounding that incorporate information from the negative control; describe a related Bayesian formulation (Appendix 2); and, provide R code to implement these analyses. Here, we measure effects with risk ratios observable in a cohort study. These approaches rely on substantive knowledge to inform the choice of the prior distribution of plausible bias parameters.

We assume measured confounders, denoted collectively by *X*, are categorical, or can be adequately approximated as categorical to control confounding (this imposes little restriction other than regularity conditions). All results are conditional on *X*, but for simplicity that dependence is suppressed in the notation. For example, *M*_*enx*_ denotes the number of people at baseline in the cohort with *E* = *e, N* = *n, X* = *x* for *e, n* = 0,1 and *x* = 1,2, …, |*X*| where |*X*| is the cardinality of *X*; but for simplicity we write *M*_*en*_ (conditioning on *X* = *x* is implicit). Conditional risk, defined as the probability that the outcome occurs (*Y* = 1) during the follow-up period among those with *E* = *e, N* = *n* at baseline in the cohort, is denoted by *R*_*en*_ = *p*(*Y* = 1|*E* = *e, N* = *n*) = *E*(*Y*|*E* = *e, N* = *n*). We denote the counterfactual outcome and counterfactual risk among those with *E* = *e, N* = *n*, if *E* were set to *e*′ by *Y*(*e*′) and *R*_*en*_(*e*′) = *E*[*Y*(*e*′)|*E* = 1, *N* = *n*] for *e, n, e*′ = 0 or 1 (Table 1), respectively; *R*_*en*_(1) or *R*_*en*_(0) must be counterfactual. Similarly, *R*(*e*) is the counterfactual risk in the population if *E* were set to *e* for all.

**Table 1.**
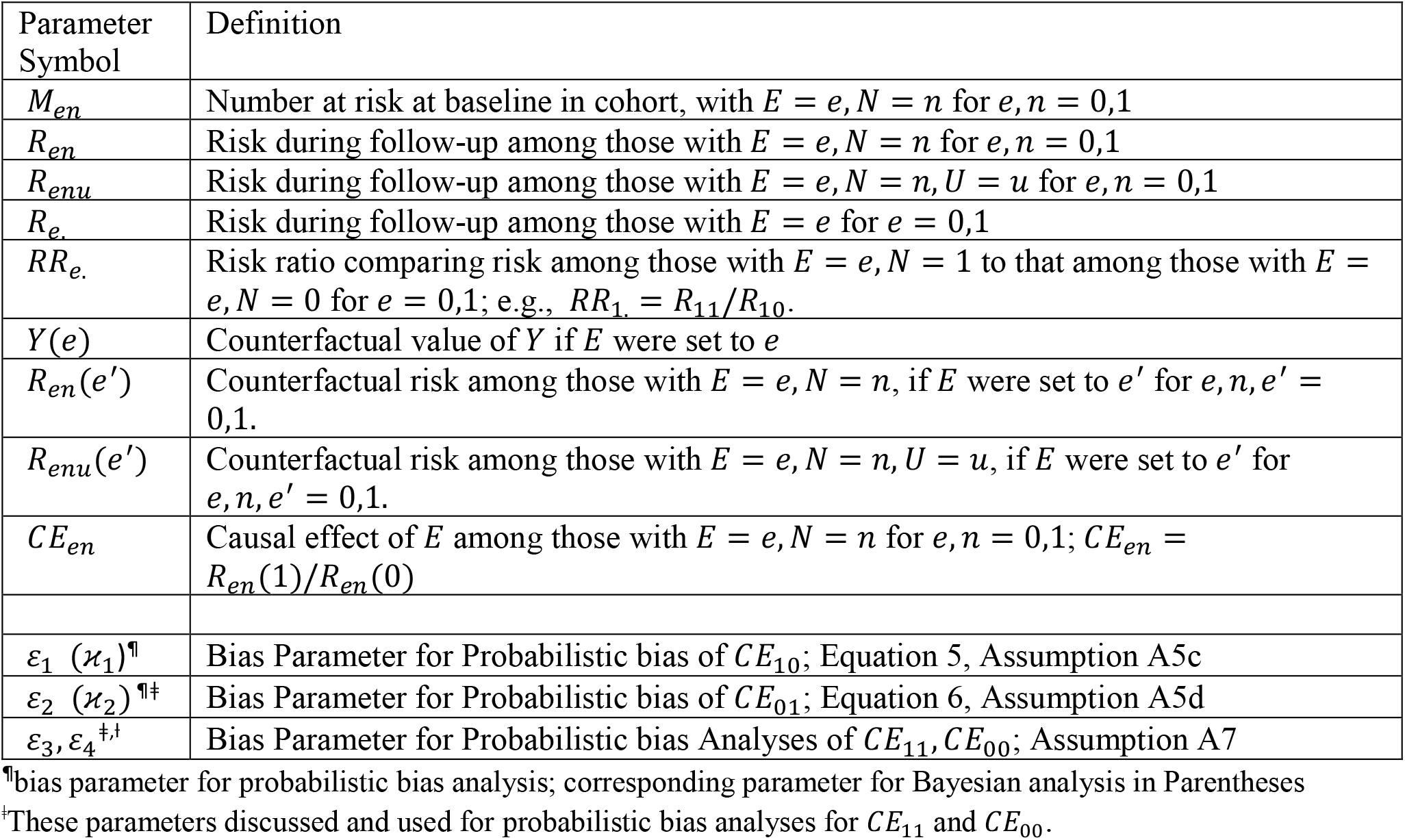
Summary of Parameter Definitions and relationship to Causal Effects

The assumed causal relationships are summarized in Figure 1, a Single World Intervention Template (SWIT)^18^. The causal relationships in the SWIT, assumed correct, imply and are consistent with assumptions A1–A4:

**Figure 1.**
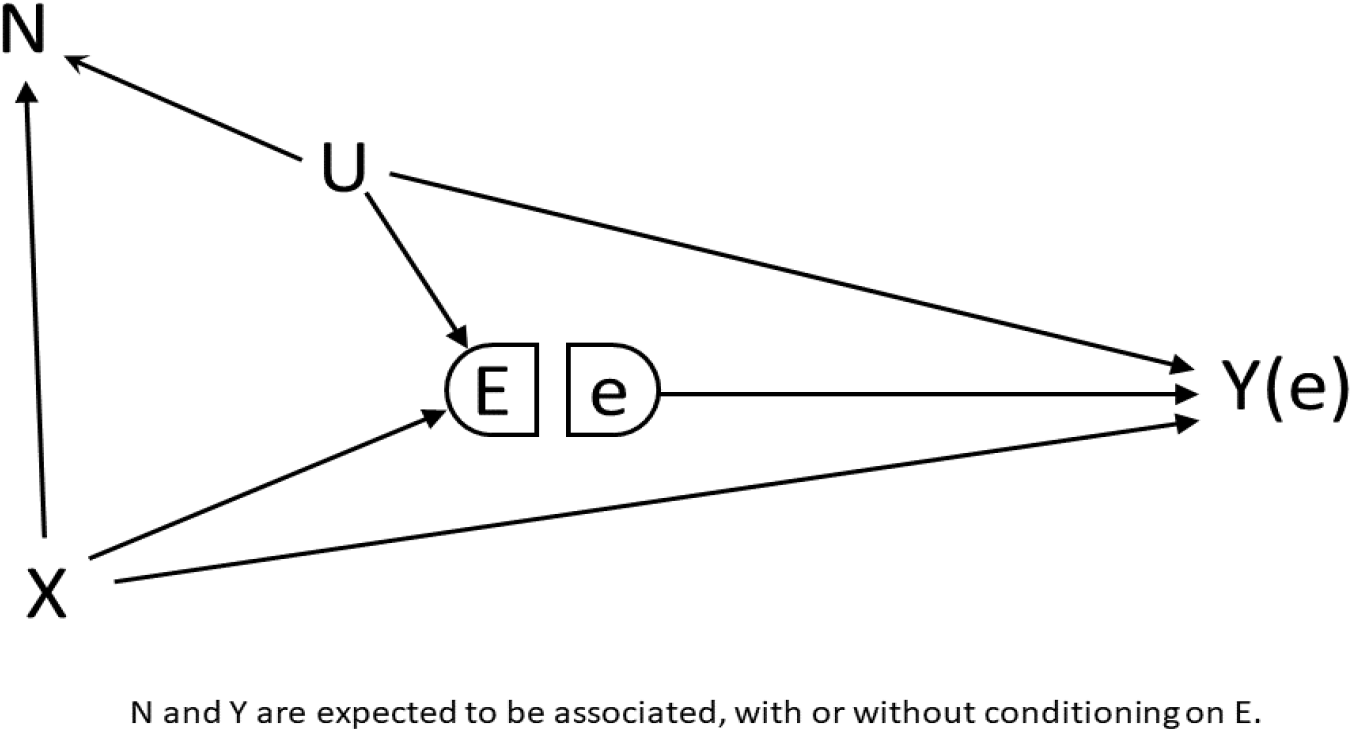
Single World Intervention Template (SWIT)^18^ showing causal relationships. *E* represents the exposure variable, *X* measured confounder(s), *U* unmeasured confounder(s), *N* a negative control and *Y*(*e*) the potential outcome of *Y* if *E* were set to *e*. These relationships are assumed correct, and are consistent with assumptions A1–A4.

A1) *N* ∐ *E, Y*(*e*) |*U, X*; *Y*(*n, e*) = *Y*(*e*), conditional independence between *N* and *E, Y*; *N* has no effect;
A2) *E* ∐ *Y*(*e*) |*U, X*; conditional exchangeability;
A3) Conditional on *E, X*, we expect the negative control to be associated with *Y* and unmeasured confounders *U*;
A4) *R*_*en*_(*e*′) = *R*_*en*_ and *R*_*en,u*_(*e*′) = *R*_*en,u*_ if *e*′ = *e*; counterfactual-model consistency.

Ideal or U-comparable negative control exposures described by Lipsitch et al.^8^ should satisfy assumptions A1 – A4 (Web Appendix S4). However, additional variables that are not ideal or U-comparable negative controls can satisfy A1 – A4, serve as indicators of residual confounding or other bias^10^ and be used for the probabilistic bias or Bayesian analyses described here (e.g., Supplemental Figure S3).

We consider the causal effects of exposure among those with *E* = *e, N* = *n*, denoted by *CE*_*en*_ for *e, n* = 0,1 (Table 1). Using risk ratios and counterfactuals, we express *CE*_*en*_ as:

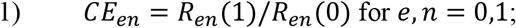

and, the population average causal effect as:

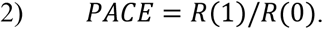

In the remainder of methods we first consider *CE*_10_ in detail, providing and justifying assumptions under which *CE*_10_ can be identified. We then introduce a bias parameter to relax the assumption needed for identification and use this parameter as the basis for probabilistic bias analyses. Finally, we consider other causal effects. In Appendix 2 provide a fully Bayesian formulation of our approach.

### *Identifiability Conditions and Probabilistic Bias Analysis for CE*_10_

We show that *CE*_10_ is identifiable in a cohort study under an easily-specified, but strong assumption involving the distribution of the negative control *N*.

Consistent with the pattern of causal effects summarized in Figure 1 and assumptions (A1)-(A4), we can write the identifiable risk *R*_00_ as:

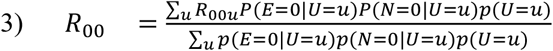

where *R*_*enu*_ is the conditional risk among those with *E* = *e, N* = *n, U* = *u*. Let *R*_*enu*_(*e*′) denote the counterfactual risk among those with *E* = *e, N* = *n, U* = *u* and *E* was set to *e*′. We can write the counterfactual risk *R*_10_(0) as the weighted average:

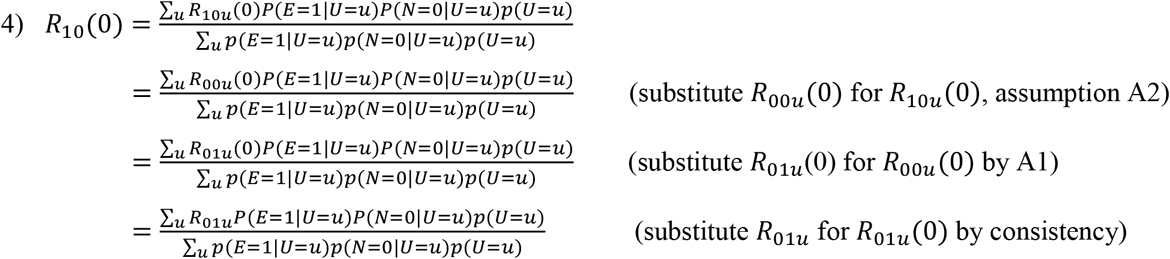

We now state two assumptions either of which, with Assumptions (A1-A4), suffices (Claim 1) to assure identifiability of *CE*_10_:

A5a) *p*(*E* = 1|*U* = *u*) = *p*(*N* = 1|*U* = *u*) for all *u*; (equality of conditional distributions) or
A5b) *p*(*U* = *u*|*E* = 1, *N* = 0) = *p*(*U* = *u*|*E* = 0, *N* = 1), for all *u*.

#### Claim 1

Under assumptions (A1-A4) and (A5a) or (A5b): (*i*) *R*_10_(0) = *R*_01_; and (*ii*) *CE*_10_ is identified by the ratio of observable risks *R*_10_/*R*_01_.

<u>Proof</u>: By assumption (A5a), we can substitute *p*(*N* = 1|*U* = *u*) for *p*(*E* = 1|*U* = *u*), and *p*(*E* = 0|*U* = *u*) for *p*(*N* = 0|*U* = *u*) into the last line of Expression (4), showing that 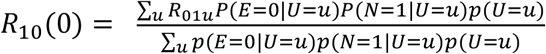 which equals *R*_01_ proving (*i*). Proof of Claim (*i*) using (A5b) is similar (Web Appendix S5). Now, 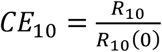 which, by (*i*), equals 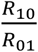. The latter is consistently estimated by the ratio of observable risks in the appropriate subgroups of the cohort study, proving (*ii*). Following Claim 1, we take the identifiable risk ratio *R*_10_/*R*_01_ as the estimator of *CE*_10_.

Note: The intuition behind this estimator is that the distortion caused by the association of the unmeasured variable U with exposure – is compensated for and balanced by the association of U with the negative control; under assumption (A5b), the distribution of U is the same in the groups being compared.

Assumptions (A5a-A5b) differ from the equi-distributional confounding assumption of Sofer et al.^19^ which concerns equality of the conditional distributions of the outcome and of a negative control outcome (NCO); see also the “confounding bridge” assumption of Miao et al.^20^ that involves conditional distributions of the NCO and the outcome, rather than the negative control exposure and the exposure.

There is some plausibility that assumption (A5a) or (A5b) would hold, at least approximately, since negative controls “… should be selected such that they share a common confounding mechanism as the exposure and outcome variables …”^2^. Nevertheless, the assumption (A5a) or (A5b) is strong and, with *U* unmeasured, unverifiable. Therefore, we introduce a bias parameter that allows for deviations from (A5a-A5b) and that can be used in probabilistic bias analyses. In particular, we relax the key implication of assumption (A5a, A5b) that *R*_10_(0) = *R*_01_, and instead assume:

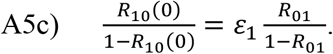

A5c assumes that the counterfactual odds *R*_10_(0) is equal to the (observable) odds *R*_01_/(1 − *R*_01_) times a bias parameter *ε*_1_. Using risk odds in assumption (A5c), rather than say risks, assures that *R*_10_(0) is not outside the range (0,1) for *ε*_1_*ϵ*(0, ∞). Substituting *ε*_1_*R*_01_/(1 − *R*_01_ + *ε*_1_*R*_01_) for *R*_10_(0), justified by Assumption (A5c), gives:

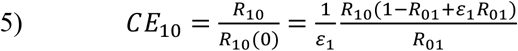

Note: The bias parameter *ε*_1_ reflects residual bias in *R*_10_ as an estimator of the counterfactual risk *R*_10_(0) on the odds scale. Equation (5) shows that *ε*_1_ equals the ratio of the estimator to the causal effect, sometimes referred to as the confounding risk ratio^1,21-23^, multiplied by (1 − *R*_01_(1 − *ε*_1_)). For rare outcomes, (1 − *R*_01_(1 − *ε*_1_)) ≈ 1, and *ε*_1_ approximates the confounding risk ratio. This conceptualization may aid interpretation (see also, Supplemental Web Appendix S1).

To implement a probabilistic bias analysis for residual confounding, we specify a distribution for *ε*_1_ (with support from 0 to infinity); the distribution has greater or less weight in the tails, depending on the extent to which *R*_10_(0) and *R*_01_ are thought to differ (reflecting differences in the conditional distributions of *E* and *N*). If the negative-control association with *U* is thought to mirror the corresponding exposure association fairly accurately, the *ε*_1_-distribution can be formulated with a substantial probability that *ε*_1_ is near 1, whereas if the associations differ substantially, greater weight can be assigned elsewhere. An R program to implement probabilistic bias analysis is given in Web Appendix S2. Using a Monte Carlo approach, the program: randomly selects a value of the bias parameter from the specified distribution; applies the bias model to calculate the counterfactual risk (*R*_10_(0), Assumption A5c); accounts for random error by sampling *R*_10_(0) and *R*_10_ from binomial distributions; applies Equation (5) to calculate a bias-adjusted estimate of *CE*_10_; and creates a simulation interval.^17^

### *Identifiability Conditions and Probabilistic Bias Analysis for CE*_01_

Assumptions (A1-A4; A5a or A5b) imply that *CE*_01_ = *CE*_10_ (proven as Claim 2 in Web Appendix S5), which implies that *CE*_01_, like *CE*_10_, is identifiable as the ratio of observable risks *R*_10_/*R*_01_. To relax assumption (A5a or A5b) and conduct probabilistic bias analyses, we introduce a second bias parameter (*ε*_2_), that plays a role like that of *ε*_1_ : *ε*_2_ reflects differences between the estimator 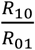 based on observed risks and the estimand *CE*_01_. We have assumption (A5d):

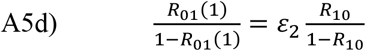

implying:

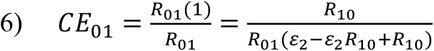

By specifying a distribution for *ε*_2_ we can conduct probabilistic bias analyses for *CE*_01_, like those for *CE*_10_.

### *Identifiability Conditions and Probabilistic Bias Analysis for CE*_11_, *CE*_00_ and *PACE*

Causal effects *CE*_11_, *CE*_00_ and *PACE* can differ from *CE*_10_ and *CE*_01_ and so are not necessarily identified as *R*_10_/*R*_01_ under Assumptions A1-A4, A5b. However, we can identify these effects if we can assume a multiplicative model for the effect of *E* given *U*, conditional on *U* = *u* and *N* = *n*:

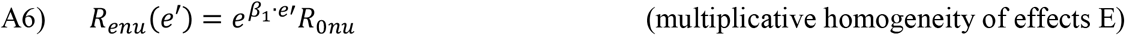

In words, assumption (A6) states that the counterfactual risk, if *E* were set to *e*′, is 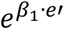 times the risk among those with *E* = 0, *N* = *n* and *U* = *u*. We show in Appendix 1 (Claim 3) that Assumption (A6) implies that *CE*_*en*_ and *PACE* both equal 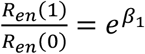. By Assumptions (A1-A5), *CE*_10_ (and *CE*_01_) are identified, and therefore so are *CE*_11_,*CE*_00_ and *PACE*. Using bias parameters to account for errors in assumptions (A1-A6) and combining results, we have

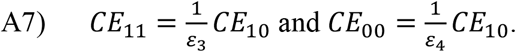

Analyses based on use of *ε*_3_ and *ε*_4_ use the strong assumption of a multiplicative effect of exposure in addition to (A1-A5) and are viewed as supplementary.

### *Fully Bayesian Analysis for CE*_10_ and *CE*_01_

Appendix 2 outlines use of prior distributions for *R*_10_ and *R*_01_ and two parameters *x*_1_ and *x*_2_ to provide a fully Bayesian formulation of the problem^24^. Parameters *x*_1_ and *x*_2_ (defined in Appendix 2: Equation 1A and just below) reflect the same associations in the Bayesian formulation as do *ε*_1_ and *ε*_2_ in probabilistic bias analyses. Web Appendix S3 documents an R program to implement Bayesian analyses for *CE*_10_ and *CE*_01_.

## Example

We illustrate these methods by applying them to investigate the possible effect of gender-affirming hormone therapy on risk of suicide attempts. We use data from ongoing studies of transgender people ^25,26^, summarized in Table 2. The cohort consists of people from two health plans in California, who were 20 years old or younger on December 31, 2015, and received a transgender-specific diagnosis (e.g., ‘gender dysphoria’) by age 20. We defined exposure as receiving gender-affirming hormones or puberty suppression therapy at or before age 20. The outcome of interest was at least one episode of self-inflicted injury or poisoning, or any hospitalization or emergency room visit for a mental health problem documented in the medical records during the 1-year follow-up period starting at age 20. Web Appendix S6 includes additional descriptive information about the cohort. We were concerned about potential confounding by healthcare utilization, as greater utilization might associate with both more hormone therapy and more (documentation of) mental health diagnoses. Therefore, we used recorded receipt of TdaP vaccine at or before age 20 years as a negative control exposure. If healthcare utilization was a confounder, we thought that TdaP vaccination should, like hormone therapy, be associated with both healthcare utilization and the outcome. The crude risk ratio (*cRR)* for exposure is 0.88. After adjusting for the negative control, the Mantel-Haenszel *mRR* is 0.88 (95% CI: 0.56 – 1.38; Table 3). The negative control was associated with risk, both among the exposed (*RR*_1_ = 1.70; 95% *CI*: 0.76 − 3.79) and the unexposed (*RR*_2_ = 1.66; 95% *CI*: 1.07 − 2.56).

**Table 2.** Distribution of Self-harm Episodes (*Y*), Hormone Use (*E*), Prior Vaccination (*N*)

| Variable | - Value of the Variable - |  |  |  |  |  |  |  |
| --- | --- | --- | --- | --- | --- | --- | --- | --- |
| Number self-harm episodes ($Y$) | 1+ | 0 | 1+ | 0 | 1+ | 0 | 1+ | 0 |
| Hormone use ($E$; yes/no) | yes | yes | no | no | yes | yes | no | no |
| Vaccinated ($N$; yes/no) | yes | yes | yes | yes | no | no | no | no |
| Number with this combination | 10 | 58 | 30 | 152 | 11 | 116 | 42 | 380 |

**Table 3.**
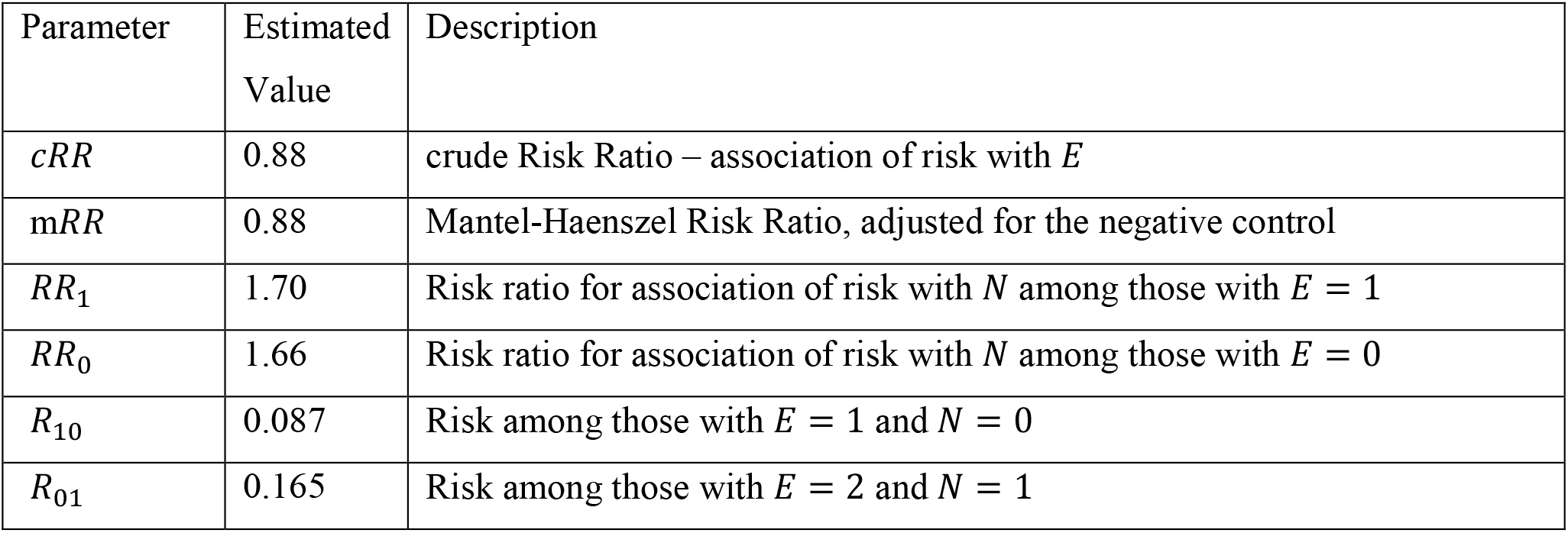
Summary of Estimated Values of Selected, Identifiable Parameters in the Example

To implement the probabilistic bias analyses, we specify prior distributions for *ε*_1_. As provided in the R program (Web Appendix S2), we chose a log-normal distribution for *ε*_1_, with median 1 and the ratio of the 10^th^ percentile to the median of 0.5. For this specification, the 10^th^ and 90^th^ percentiles of the prior for *ε*_1_ are 0.50 and 2.0, indicating that *ε*_1_ will fall in this range with 80% probability under the prior. The resulting distribution of confounding-adjusted causal effect estimates (simulation intervals)^17^ is given in Figure 2, for the assumed prior. The 95% simulation interval is from 0.17 to 1.64, with median 0.52. These results are approximately interpretable as semi-Bayesian, as the bias parameter is sampled from a prior distribution^17^. Using the fully Bayesian analysis described in Appendix 2 with uninformative priors for *P*(*Y* = 1|*E* = 1, *N* = 2) and *P*(*Y* = 1|*E* = 2, *N* = 1), a log-normal prior for *x*_1_) and setting the median of the prior (log-normal) to 1 and the variance so that the ratio of the median to the 10^th^ percentile was 0.5, the 95% credibility interval was (0.19 to 1.69) with median 0.54. In a sensitivity analysis, we doubled the variance of the prior for *κ*_1_-reflecting greater uncertainty in the value of the bias parameter. The 95% credibility interval was then (0.16, 2.08).

**Figure 2.**
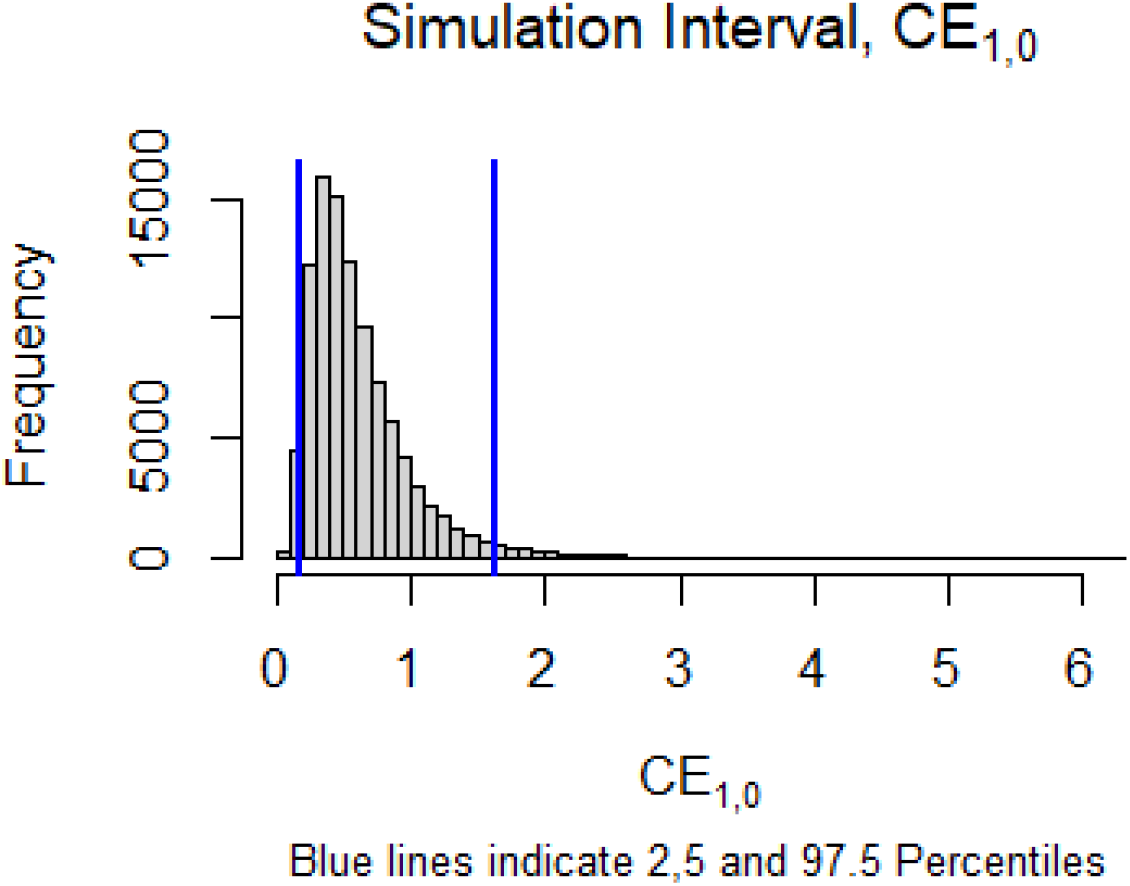
Histogram showing the distribution of confounding-adjusted causal effect estimates from probabilistic bias analysis, with the prior distribution of the bias parameter as described for the Example.

## Discussion

We have justified assumptions that, if correct, imply one can use a negative control exposure to account for unmeasured confounding and identify causal effects (*CE*_10_ and *CE*_01_). If exposure effects are multiplicative, this suffices to also identify *CE*_11_ and *CE*_00_ as well as other effects, such as the population average effect. The key assumption (A5a or A5b) is strong, so we have also described and illustrated two ways to relax the assumption. These inter-related methods, probabilistic bias analyses and Bayesian analyses, both use information from a negative control exposure to account for residual confounding and require the researcher to specify a prior distribution for a bias parameter; they yielded similar results, as expected, in our example. Our formulation of probabilistic bias analyses, as is common^17^, includes a bias model, postulating a prior distribution for the bias parameters, and Monte Carlo simulation to obtain a distribution for the bias-adjusted estimate of interest. Our method extends the approach to incorporate information from the negative control. We also describe a complementary Bayesian formulation.

Exchangeability implies that risk among those actually exposed is the same as the risk among the unexposed if they had been exposed, and conversely; it can be defined as independence of the actual exposure and response types^27-29^.The methods proposed for addressing non-exchangeability are natural ones in the sense that they use the negative control as a reflection of exposure associations with unmeasured confounders that define non-exchangeability. If the negative control has more than two categories, say *N* = *n* for *n* = 0,1, …, *N*, then the approach described here is still applicable by selecting two categories (or combinations of categories) and contrasting them. For example, if a priori knowledge suggested that *P*(*U* = *u*|*E* = 1, *N* = 0) ≈ *P*(*U* = *u*|*E* = 0, *N* ∈ *S*_1_) where *S*_1_ = {3,4}, then *R*_10_(0) could be estimated by 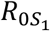. To the extent allowed by *a priori* knowledge, original categories can be combined to most closely approximate the identifying assumption.

Two causal effects are most readily addressed by the proposed approach, the effect of exposure among the exposed without negative control exposure (*E* = 1, *N* = 0), and the effect of exposure among the unexposed with negative control exposure (*E* = 0, *N* = 1). The researcher can use substantive knowledge to assess how well the association of the negative control with the unmeasured confounder reflects the association of exposure with the confounder. In the example when using Bayesian analyses, doubling the variance of the prior distribution for *κ*_1_ led to wider credibility intervals, but not substantially so (Example 1) – suggesting some degree of robustness to a modest change in uncertainty regarding the prior distribution of *x*_1_. While it is possible to extend the approach to apply it to effects in other subgroups and to a population average causal effect (Appendix 1), assignment of the prior distribution is perhaps more uncertain because an additional assumption (multiplicative effect of exposure) is needed. Therefore, we view these additional analyses as secondary. We caution that an association between a negative control and the outcome can reflect bias other than non-exchangeability, such model mis-specification^5^. Our analyses are not designed to correct for these other biases.

Results of the probabilistic bias analysis and the Bayesian analysis both utilize a negative control to correct for residual confounding and are based on researcher-supplied inputs that are informed, to the extent possible, on subject matter knowledge. The probabilistic bias analyses depend on the prior distributions for *ε*_1_ and *ε*_2_ and the Bayesian results on those for *x*_1_ and *x*_2_; here we used log-normal distributions for both. With a non-informative prior for the other parameters, the 95% credibility interval (Bayesian analysis) was similar to the simulation interval (probabilistic bias analysis). Absent assumptions (such as A5b), parameters *x*_1_ and *x*_2_ or bias parameters *ε*_1_ and *ε*_2_ are not identifiable; however, “indirect learning” is possible^30^, evidenced here in the change from prior to posterior distribution of *κ*_1_(Web Appendix S1: Figures S1 and S2). This indirect learning and changes in the distributions result from learning about the identifiable parameters^30^.

In some situations, when the confounder is known but unmeasured, external results may provide direct estimates of *κ*_1_ or *ε*_1_. For example using equation 5, *ε*_1_ could be estimated as 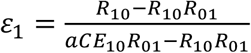 where *aCE*_10_ is an external estimate of the causal effect (e.g., adjusted for all confounders in a study where U was measured). However, if external measurements of the exposure, confounders and outcome are available, we can also consider other, possibly more efficient, approaches^17^ or perhaps use of a directly calculated confounding risk ratio. We could also use priors for both *κ*_1_and a causal effect (e.g., *CE*_01_), plus another parameter (e.g., *R*_10_). Evaluation of the posterior distribution, however, would likely then require Gibbs sampling or other technique more complicated than the straightforward one used here.

In summary, we have provided assumptions sufficient for using a negative control to identify causal effects when a confounder is unmeasured, and have described and illustrated the application of both probabilistic bias analysis and Bayesian formulations to address residual confounding. The latter methods use a negative control exposure, use researcher-supplied prior information about how well the negative control captures the associations that create confounding, and produce results partially adjusted for the residual confounding.

## Data Availability

Only the summary data in Table 2 are needed to replicate the analysis.

## Abbreviations

RR: risk ratio
CE: causal effect.

## Appendix 1

### Claim 3

Under assumptions (A1-A4, A5b, and A6), 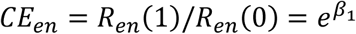 and 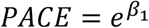.

<u>Proof</u>: *R*_*en*_ (*e*′) is a weighted average of the counterfactual outcomes *R*_*enu*_(*e*′). Therefore:

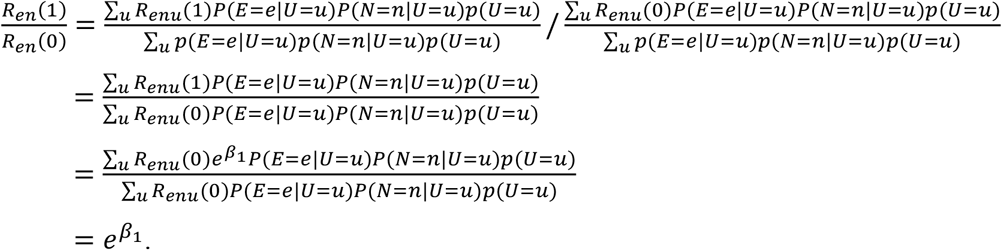

Thus: *CE*_11_ = *CE*_00_ = *CE*_10_ = *CE*_01_. The *PACE* is a weighted average of the four *E, N* – specific effects *CE*_*en*_ (with weights *P*(*E* = *e, N* = *n*)), so *PACE* equals the common value 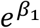.

## Appendix 2

The Appendix describes a Bayesian formulation of our approach. The analytic goal now is to calculate Bayesian credibility intervals and the posterior median for the causal effects, conditional on the observations and using the negative control exposure. We use the terminology and definitions from Lash^1,17^. We present details only for the analysis of *CE*_10_, as those for *CE*_01_ are directly analogous. Results for *CE*_11_ and *CE*_00_ depend on an additional, strong assumption and are considered supplementary.

### Parameters

The bias parameter *ε*_1_ of the main text was introduced to relax assumption (A5a); it allows for differences between the distribution of *E* given *U* and that of *N* given *U*. Following assumption (A5c), we define the (relative) bias parameter *x*_1_ as:

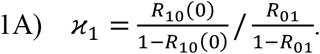

Parameters *R*_10_, *R*_01_ (Table 1) and *κ*_1_ fully parameterize the conditional distributions of: outcome *Y* and *Y*(0) among those with *E* = 1, *N* = 0, and those among individuals with *E* = 0, *N* = 1. For example, the distribution of *Y*(0) among those with *E* = 1, *N* = 0 is:

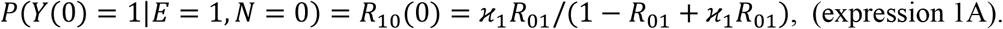

*CE*_10_ can be expressed using only *R*_10_, *R*_10_ and *x*_1_. Much like *ε*_1_, we define 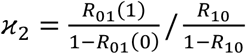 so that *P*(*Y*(1) = 1|*E* = 0, *N* = 1) = *R*_01_(1) = *R*_10_/*R*_01_(*ε*_2_ − *ε*_2_*R*_10_ + *R*_10_). This formulation parallels that of probabilistic bias analysis in the main text; e.g., Equation 1A replaces Assumption A5a).

### Sampling Distribution

The conditional likelihood of the observed data, given the parameters *R*_10_ = *p*_1_ and *R*_01_ = *p*_2_ is:

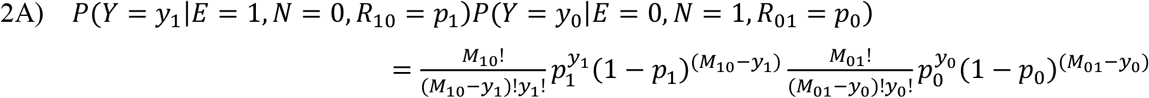

*y*_1_ is the number of subjects with *y*_1_ = 1, *E* = 1 and *N* = 0; *M*_10_ is the number with *E* = 1 and *N* = 0; *y*_0_ and *M*_01_ are the corresponding numbers where *E* = 0 and *N* = 1; the “data” are *y*_1_, *M*_10_, *y*_0_, *N*_01_, *E* and *N*.

<u>Prior Distributions</u> for parameters *R*_10_, *R*_01_, and *x*_1_

We use a log-normal prior for *x*_1_:

3A) 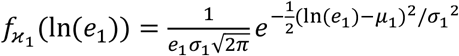 for 0 ≤ *x*_1_ < ∞ and 0 elsewhere, where *µ*_1_ and *σ*_1_^2^ are the mean and variance of ln(*e*_1_) The median of *e*_1_ is 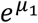 and the variance is 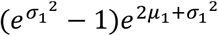. To double the variance of *e*_1_ (on the original, non-log scale), we solve: 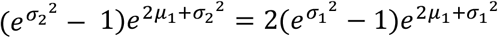 for *σ*_2_ for. For *σ*_1_ = 0.54 (the initial value of in *σ*_1_ used in the Example, main text), we use *σ*_2_ = 0.67 to double the variance of *e*_1_.

We use a beta prior for *R*_10_:

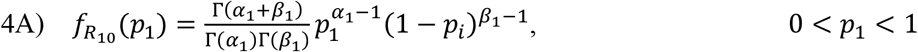

and a beta prior for *R*_01_:

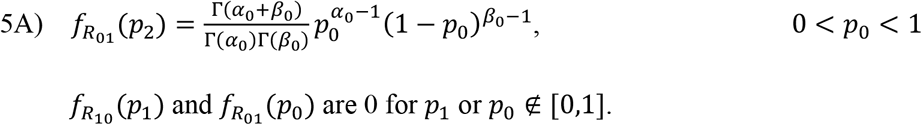

The priors 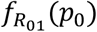 and 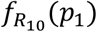 are non-informative uniform priors if we use *α*_*j*_ = *β*_*j*_ = 1.

### Posterior Distribution

For analysis of *CE*_10_, the posterior distribution is:

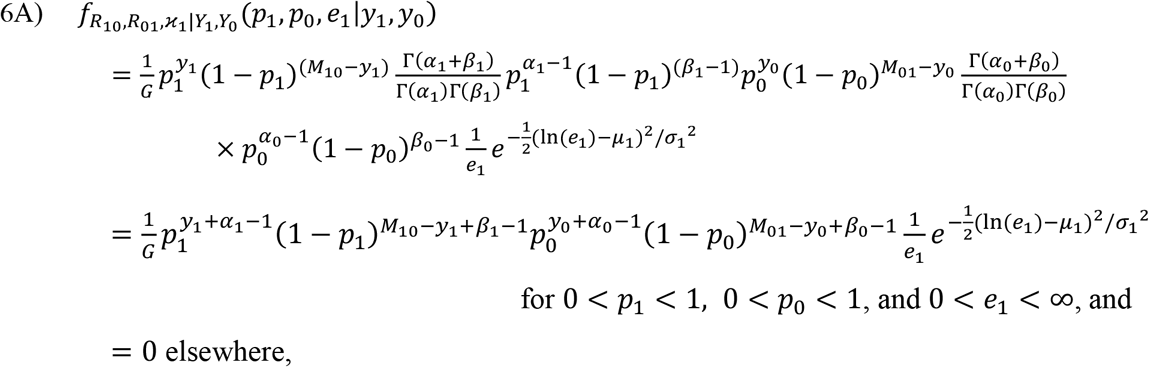

Where 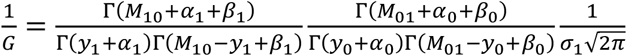.

The posterior distribution for analysis of *CE*_10_ is the product of two independent *beta* distributions and a *log-normal* distribution. It is therefore straightforward to sample from this joint distribution, by independently sampling: *p*_1_ (for *R*_12_) from a *beta*(*y*_1_ + *α*_1_, *M*_10_ − *y*_1_ + *β*_1_), *p*_0_ (for *R*_01_) from *beta*(*y*_0_ + *α*_0_, *M*_01_ − *y*_0_ + *β*_0_), and *e*_1_ (for *κ*_1_) from a *log-normal(µ*_1_, *σ*_1_^2^) distribution.

### Evaluation

To evaluate the posterior distribution, the supplemental R code (Web Appendix S3) performs the described sampling in 100,000 independent replications. For each sample, it calculates 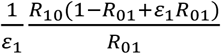 (or 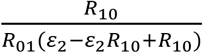), equal to the parameter of interest *CE*_10_ (or *CE*_10_). It then calculates desired statistics from the empiric distribution of sampled values (e.g., 2.5 and 97.5 percentiles for the 95% credibility interval for *CE*_10_, and the 50^th^ percentile for the median). The program uses *α*_*i*_ = *β*_*i*_ = 1 as the default parameters for the *beta* distribution, as this choice yields uninformative, uniform priors. The user must input the parameters of the log-normal prior distribution of *x*_*i*_ (details and further explication in Web Appendices S1-S2).

## Online Supplement

### Web Appendix S1

This Web Appendix (S1) includes additional comments about use of the R programs (Web Appendix S2 and S3).

Probabilistic bias analyses (R program in Appendix S2) require the user to specify a distribution for *ε*_1_ (or *ε*_2_). Specifying a distribution with a median of 1 (e.g., as in the example, main text), would be appropriate if *a priori* information suggested that the counterfactual odds 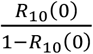 is expected to exceed the true, identifiable odds 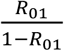 with about 50-50 probability. However, if *a priori* information suggests that the counterfactual odds is more likely than not to be larger than the identifiable odds 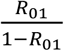 or,similarly, that *R*_10_/*R*_01_ is likely to over-estimate the causal effect *CE*_10_, then the median should be chosen greater than 1. This might reflect a situation in which exposure was more strongly associated with high-risk values of U than was the negative control. Conversely, if the counterfactual odds is more likely than not to be smaller than the identifiable odds 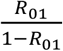, then the median should be chosen less than 1. The variance reflects uncertainty, with larger variances indicating a wider range of values of *ε*_1_ judged to be compatible with a priori information.

Bayesian analyses require the user to specify the prior distribution of *κ*_1_ (corresponding comments also hold for *κ*_2_). To do so using the R program supplied (Appendix S3), the user inputs the median of *x*_1_, which is the mode of the *log-normal* distribution used as the prior for *κ*_1_. The log-normal parameter *µ*_1_ (equation 6A of the Appendix, main text) is the logarithm of this median. The user also inputs the desired value for the ratio of the 10^th^ percentile of *κ*_1_ divided by the median in the distribution of *κ*_1_. The program uses these inputs to calculate the variance *σ*_1_^2^(equation 6A of the Appendix 2, main text) that yields these user-supplied percentiles. It then generates random samples from the posterior distribution (that depends on the parameters as described. To illustrate, suppose, for example, that the researcher thought that the median was 1 and that the 10^th^ percentile was one-half as large as the median. This relationship would hold if the standard deviation of the log-normal prior were set to 0.54. In this case, the prior would specify that 80% of the values of *κ*_1_ were between 0.5 (the 10 percentile) and 2.0 (the 90 percentile). Since 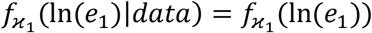, i.e. the posterior and prior are the same for *κ*_1_, we expect the Bayesian analysis to agree closely with the probabilistic bias analysis.

Note S1: The analysis described for *CE*_10_ (or *CE*_01_) is fully Bayesian as all parameters needed to write the (conditional) probability of the data used have a prior, a reflection of the relationships in equations 2A-6A, Appendix 2 of the main text. In particular, a prior distribution for the causal effects *CE*_10_ (or *CE*_01_) is indirectly specified through the priors for *R*_10_, *R*_01_ and *x*_1_ (or *x*_2_).

Note S2: To measure causal effects using risk odds ratios (rather than risk ratios as above), then we can define 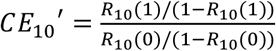. The definitions (of *R*_10_, *R*_01_, *κ*_1_ and *CE*_10_) and distributions above are unchanged (programs to conduct probabilistic bias and Bayesian analyses using risk ratios are given in Web Appendices S2 and S3).

Comparison of the prior distribution for *κ*_1_, illustrated by empirically simulating it (Figure S1) for the situation described in the example (main text), with the corresponding posterior distribution (Figure S2) shows a meaningful shift (e.g. median changes from 0.99 to 0.48), illustrating that the data do provide information about the causal effect under the assumed priors.

Note S3: Specification of priors for *R*_10_, *R*_01_ and *κ*_1_ implies a prior for *CE*_10_ because of the deterministic relationships (equation 5, main text). In view of these inter-relationships of the parameters, specifying a prior for the causal effects themselves, e.g. for *CE*_10_, in addition to priors for *R*_10_, *R*_01_ and *κ*_1_ could lead to inconsistencies in view of the deterministic relationships. However, one could specify priors for, say, *R*_10_, *x*_1_ and *CE*_10_, but not, say, *R*_01_. In this case the posterior distribution would be different from that in Appendix 2 (main text), but could be evaluated using Gibbs sampling or other Markov Chain Monte Carlo approach. Here, the approach implemented in the program specifies a prior for the causal effect (*CE*_10_) indirectly through those for *R*_10_, *R*_01_ and *κ*_1_ (Figure S1; the prior for example of main text), and does not separately specify a prior for *CE*_10_.

Note S4: If direct evidence, *a priori* considerations or otherwise expected differences between the *E* and *N* conditional distributions (departures from assumption A5) suggest that *R*_10_(0) will tend to exceed *R*_01_ so that *κ*_1_is less than 1 (in Assumption A5a), that would suggest that *E* tends to more strongly associate with high-risk values of *U* than does *N*. This pattern suggests that *R*_01_(1) should tend to be less than *R*_10_ and that *κ*_2_ may be greater than 1. These considerations suggest, that if the prior distribution of *x*_1_ has substantial mass below 1, then that for *κ*_2_ might reasonably specified to have substantial mass above 1 (and conversely).

### Web Appendix S2

Appendix S2 is an R program for Probabilistic Bias Analysis; effects are measured using risk ratios (See also Web Appendix S1).

~~~
# Probabilistic Bias Analysis for Causal Effect CE21 (or CE12), using Risk Ratios
library(tidyverse)
library(pscl)
library(dplyr)
library(data.table)
library(readxl)
# ------------- user inputs ---------------------------- #
med = 1  #user input, desired median (log(med)=mean yields mu, of log-normal dist)
q10 = 0.5  #user input, desired ratio: 10-percentile/med: used to calculate sd
n=100000  #user input, number of random draws
N = 1

## ------------------------ input data ----------------------------------
dat <- array(rep(0,8), dim=c(2,2,2))
dat[1,1,1]= 10 #P(Y=1, E=1, NC=1)
dat[2,1,1]= 58
dat[1,2,1]= 30
dat[2,2,1]= 152

dat[1,1,2]= 11 #P(Y=1,E=1,NC=2)
dat[2,1,2]= 116
dat[1,2,2]= 42
dat[2,2,2]= 380

n1 = dat[1,1,2]+dat[2,1,2]
n2 = dat[1,2,1]+dat[2,2,1]

## --------------------------- END: input data ------------------------------------

save = NULL

# ----- calculate conditional Probs from input dat, so they to sum to 1
for (j in 1:2){
 for (k in 1:2){
  dat[1,j,k]=dat[1,j,k]/sum(dat[,j,k])
  dat[2,j,k]=1-dat[1,j,k]
  }}
dat
# ----- END: calculate cond probs.

AVERAG<-0
use=NULL
# ===================--------------------------------------------===============
# ================== -------- Crude Risk Ratio --------- ==================
RR1= dat[1,1,1]/dat[1,1,2]
RR2= dat[1,2,1]/dat[1,2,2]
cRR = (dat[1,1,1]+dat[1,1,2])/sum(dat[,1,])
cRR = cRR/((dat[1,2,1]+dat[1,2,2])/sum(dat[,2,]))
R11= dat[1,1,1]
R12= dat[1,1,2]
R21= dat[1,2,1]
R22= dat[1,2,2]

mRR= dat[1,1,1]*(dat[1,2,1]+dat[2,2,1])/sum(dat[,,1])
mRR=mRR+ dat[1,1,2]*(dat[1,2,2]+dat[2,2,2])/sum(dat[,,2])
den= dat[1,2,1]*(dat[1,1,1]+dat[2,1,1])/sum(dat[,,1])
den=den+ dat[1,2,2]*(dat[1,1,2]+dat[2,1,2])/sum(dat[,,2])
mRR= mRR/den
mRR
#RD_E.NC #association of E, NC

crude=data.frame(RR1=RR1, RR2=RR2, R11=R11, R12=R12, R21=R21, R22=R22, cRR) #RD for
Exposure, then for NC
crude
c(R11/R12, R21/R22) #Y-assoc with N|E=1 or 2
#
# ------- start log-normal, prior for e1 ; -----------;
z10=qnorm(0.10, mean=0, sd = 1) #get z-score @ 10-percentile
sd = (log(q10)-log(med))/z10 #set sd on log scale so (log(q10)-log(med))/sd)=z10
z90=qnorm(0.90, mean=0, sd = 1)
c(z10, z90)
c(exp(log(med)+z10*sd),exp(log(med)-z10*sd))
e1= rnorm(n, log(med), sd)
#e2= rnorm(n, -log(med), sd) #use symetric value for e2, per Note S4
e1=exp(e1)
c(quantile(e1, probs=c(0.025, 0.5, 0.975)), sd) #display 95% interval for e1
#e2=exp(e2)
#c(exp(qnorm(0.10, mean=0, sd = sd)),
# exp(qnorm(0.90, mean=0, sd = sd))) #display 20%, 80% of epsilon
#reverse the order
R12_0 = (e1*R21/(1-R21))/(1+(e1*R21/(1-R21))) # calc R12(0) from R21 1st (use obs’d R21)
#r21 = rbinom(n, n2, R21)/n2   # sample R21, with random error
r12_0 = rbinom(n, n2, R12_0)/n2   # sample R12(0), with random error
r12 = rbinom(n, n1, R12)/n1   # sample R12, with random error
# in Example, v. similar result if change, sample R21 & R12 1st, correct w/e1 last
# CE12= r12*(1-r21+e1*R21)/(e1*r21)  # use this if change order, RR for CE10; modify for ROR
CE12 = r12/r12_0    # RR for CE10
#CE21= r12/(r21*(e2-e2*r12+r12))   # use this if desire RR; modify for ROR
# ---------------- Figure 2 --------------------------
hist(CE12, breaks=50, main=expression(“Simulation Interval, CE”[paste(1,”,”,0)]),
  xlab = expression(“CE”[paste(1,”,”,0)]),
  sub=“Blue lines indicate 2,5 and 97.5 Percentiles”, cex.sub = 0.8)
abline(v=quantile(CE12, probs= 0.975),col=“blue”,lwd=2)
abline(v=quantile(CE12, probs= 0.025),col=“blue”,lwd=2)
quantile(CE12, probs=c(0.025, 0.5, 0.975)) # display empiric 95% credibility interval
~~~

### Web Appendix S3

R program for Bayesian analyses, described in Appendix 2, main text (see also Web Appendix S1); effects measured using risk ratios.

~~~
# Bayesian Analysis for Causal Effect CE12 (or CE21), using Risk Ratios

library(tidyverse)
library(pscl)
library(dplyr)
library(data.table)
library(readxl)
# ------------- user inputs ---------------------------- #
med = 1  #user input, desired median (log(med)=mean yields mu, of log-normal dist)
ratio_10 = 0.5  #user input, desired ratio: 10-percentile/med: used to calculate sd
n=100000  #user input, number of random draws

## ------------------------ input data ----------------------------------
dat <- array(rep(0,8), dim=c(2,2,2))
dat[1,1,1]= 10 #P(Y=1, E=1, NC=1)
dat[2,1,1]= 58
dat[1,2,1]= 30
dat[2,2,1]= 152

dat[1,1,2]= 11 #P(Y=1,E=1,NC=2)
dat[2,1,2]= 116
dat[1,2,2]= 42
dat[2,2,2]= 380
# ========= -------- Some Risks, ORs, & RRs ------- ================
OR1= (dat[1,1,1]/dat[2,1,1])/(dat[1,1,2]/dat[2,1,2])
OR2= (dat[1,2,1]/dat[2,2,1])/(dat[1,2,2]/dat[2,2,2])

RR1= (dat[1,1,1]/dat[1,1,2])/((dat[1,1,1]+dat[2,1,1])/(dat[1,1,2]+dat[2,1,2]))
RR2= (dat[1,2,1]/dat[1,2,2])/((dat[1,2,1]+dat[2,2,1])/(dat[1,2,2]+dat[2,2,2]))
R11= dat[1,1,1]
R12= dat[1,1,2]/(dat[1,1,2]+dat[2,1,2])
R21= dat[1,2,1]/(dat[1,2,1]+dat[2,2,1])
R22= dat[1,2,2]
cRR = (dat[1,1,1]+dat[1,1,2])/((dat[1,1,1]+dat[2,1,1]+dat[1,1,2]+dat[2,1,2]))
cRR = cRR /((dat[1,2,1]+dat[1,2,2])/((dat[1,2,1]+dat[2,2,1]+dat[1,2,2]+dat[2,2,2])))
aRR = dat[1,1,1]*(dat[1,2,1]+dat[2,2,1]) + dat[1,1,2]*(dat[1,2,2]+dat[2,2,2])
aRR=aRR/(dat[1,2,1]*(dat[1,1,1]+dat[2,1,1]) + dat[1,2,2]*(dat[1,1,2]+dat[2,1,2]))

crude=data.frame(OR1=OR1, OR2=OR2, RR1=RR1, RR2=RR2, R11=R11, R12=R12, R21=R21,
R22=R22) #RD for Exposure, then for NC
(R12/(1-R12))/(R21/(1-R21))
crude
y1= dat[1,1,2]
n1= dat[2,1,2]+y1
y2= dat[1,2,1]
n2= dat[2,2,1]+y2
# -- first, sample r21 & r12
r21= rbeta(n, y2+1, n2-y2+1)
r12= rbeta(n, y1+1, n1-y1+1)
# -- second, sample e1 from log-normal prior
# use input median and ratio of median to 10th percentile: get sd
z10=qnorm(0.10, mean=0, sd = 1) #get z-score @ 10-percentile
sd = (log(ratio_10)-log(med))/z10 #set sd on log scale so (log(rtio_10)-log(med))/sd)=z10
z90=qnorm(0.90, mean=0, ratio_10, sd = 1)
c(z10, z90, med, sd)
c(exp(log(med)+z10*sd),(med),exp(log(med)-z10*sd)) #display e1 at 10%, 50%, 90%
#sd= sd3 #< ------ set sd=sd3 for sensitivity analyses to double Var of prior
     #if sd= sd3 NOT commented out, doubles Var (value from end of pgm)
#for Example, sd=0.674 <-- use to double variance from initial value of 0.54
e1= rnorm(n, log(med), sd) #sd, sd3 in a sens Anal w/double Var
c(exp(qnorm(0.10, mean=log(med), sd = sd)),
 exp(qnorm(0.90, mean=log(med), sd = sd))) #display empiric 20%, 80% of e1
e1=exp(e1)
# ------- END: sample e1 from log-normal prior -----------;

#quantile(e1, probs=c(0.025, 0.5, 0.975)) #display 2.5th 97.5 percentiles
CE12= r12*(1-r21+e1*R21)/(e1*r21) # use this for RR, could do ROR

#CE21= r12/(r21*(e2-e2*r12+r12)), for RR
# ---------------- Figure S2 ----------------------
hist(CE12, breaks=50, main=expression(“Posterior Distribution CE”[paste(1,”,”,0)]),
  xlab = expression(“CE”[paste(1,”,”,0)]),
  sub=“Blue lines indicate 95% Credibility Interval”, cex.sub = 0.8)
abline(v=quantile(CE12, probs= 0.975),col=“blue”,lwd=2)
abline(v=quantile(CE12, probs= 0.025),col=“blue”,lwd=2)
quantile(CE12, probs=c(0.025, 0.5, 0.975)) # sisplay empiric 95% credibility interval

c(R12/(R12+dat[2,1,2]), R21/(R21+dat[2,2,1]), RR1, RR2, cRR, aRR)
# -------- empirically evaluate prior for CE ------
prior=FALSE
if (prior==TRUE){
 r21= rbeta(n, 1, 1)
 r12= rbeta(n, 1, 1)
 e1= rnorm(n, log(med), sd)
 e1=exp(e1)
 s2=sd**2
 c(var(e1), (exp(s2) -1)*exp(s2))
 CE12= r12*(1-r21+e1*R21)/(e1*r21) # use this for RR, could do ROR
 quantile(CE12, probs=c(0.025, 0.5, 0.975))
 parm2 = as_tibble(CE12, CE=CE12)
 names(parm2)=“CE”
 parm2
 parm2 = subset(parm2, parm2$CE< 30)
 # ---------------- Figure S1 --------------------------
 hist(parm2$CE, breaks=50, main=c(expression(“Prior Distribution CE”[paste(“1”,”,”,”0”)]*∼”(truncated to display)”)),
  sub=“Blue lines indicate 2.5th and 97.5 percentile”, cex.sub = 0.8,
  xlab = expression(“CE”[paste(1,”,”,0)]))
 abline(v=quantile(parm2$CE, probs= 0.975),col=“blue”,lwd=2)
 abline(v=quantile(parm2$CE, probs= 0.025),col=“blue”,lwd=2)
 }
 # for use in Sensitivity (Bayesian analyses) find sd* that double var on e-scale
 #solver - arbitrary function called fx(x), one variable
 fx<- function(x){ #just an example
  s1=0.54**2 #var used (log-scale), solve for x for Var doubles (real scale)
  2*(exp(2*s1)-exp(s1)) - (exp(2*x)-exp(x)) }
sd3=sqrt(uniroot(fx,interval=c(-10,3))$root)
c(sd, sd3) #set sd=sd3 for sensitivity analyses to double Var of prior
~~~

### Web Appendix S4

Lipsictch et al^1^ note that an ideal negative control exposure would have the same causes in common with the outcome as did the actual exposure. Figure S3 is a single world intervention graph in which *E* and *N* have different causes in common with Y, but wherein *N* can still serve as a negative control exposure.

Assumption (A5), while still possible, may be now less plausible.

### Web Appendix S5

#### Claim 1

Under assumption (A1-A4) and (A5a) or (A5b): (*i*) *R*_10_(0) = *R*_01_; and (*ii*) *CE*_10_ is identified by the ratio of observable risks *R*_10_/*R*_01_.

<u>Proof</u>: Proofs of (*i*) under assumption A5a and (*ii*) are in the main text. Here we show that (A1-A4) and (A5b) implies (*i*). By conditional independence of *E* and *N* (Assumption A1) and rules of conditional probabilities:

1S) *p*(*U* = *u*|*E* = 1, *N* = 0) = *p*(*E* = 1|*U* = *u*)*p*(*N* = 0|*U* = *u*)*p*(*U* = *u*), for all *u*, and:
2S) *p*(*U* = *u*|*E* = 0, *N* = 1) = *p*(*E* = 0|*U* = *u*)*p*(*N* = 1|*U* = *u*)*p*(*U* = *u*), for all *u*. From the last line of Equation 4) of the main text:

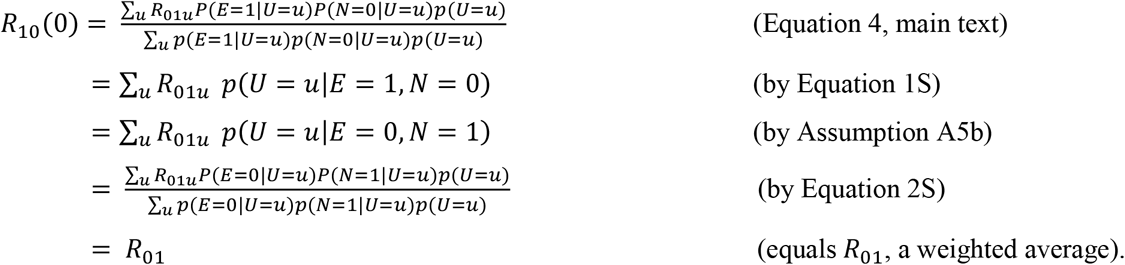

Note: Equations 1S and 2S show that Assumption (A5b) is implied by (A5a) and that (A5b) is weaker.

#### Claim 2

Under assumptions (A1-A4, A5b), *CE*_10_ = *CE*_01_.

<u>Proof</u>: The proof parallels that of Claim 1. *R*_*en*_(*e*′) is the weighted average of the counterfactual outcomes *R*_*enu*_(*e*′), weighted by *P*(*U* = *u*|*E* = *e*, N = *n*) = *P*(*E* = *e*|*U* = *u*)*P*(*N* = *n*|*U* = *u*)*p*(*U* = *u*)/∑_*u*_ *P*(*E* = *e*|*U* = *u*)*P*(*N* = *n*|*U* = *u*)*p*(*U* = *u*), so:

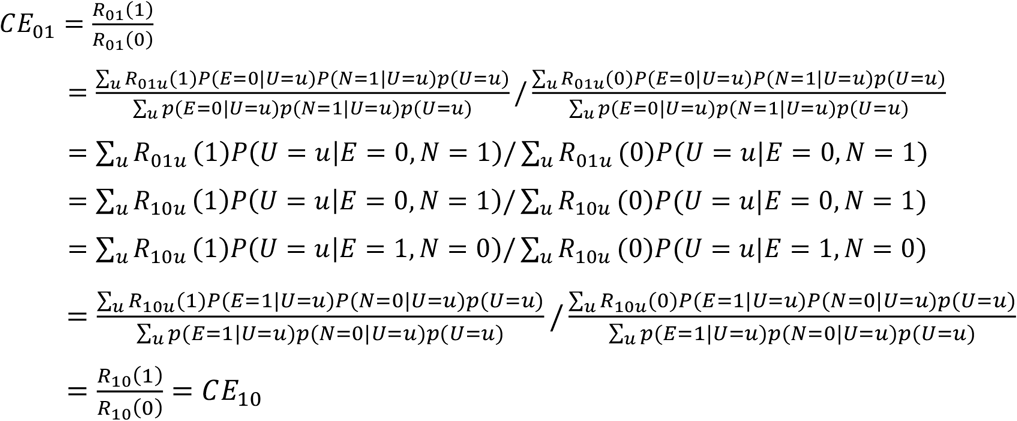

The second equality follows by properties of the counterfactuals *R*_01_(1) and *R*_01_(0), the third equality by equations 1S and 2S, the fourth by assumptions (A1) and (A2), the fifth by assumption (A5b), the sixth by equations 1S and 2S, and the last two by definitions.

### Web Appendix S6

This Web Appendix contains additional, basic descriptive information about the cohort in the Example of the main text. First, we note that, during the follow-up period, the age of cohort members with gender-affirming hormone or puberty suppression therapy, was very similar to the age of those without that therapy by definition of the cohort. In particular, the therapy had to have been received by age 20, and the follow-up for the outcome of interest was from age 20 to age 21. Thus, age during the time at risk should not have led to meaningful confounding. Table S1 includes additional descriptive information for cohort of the example in the main text and shows reasonable degree of balance between treatment groups – so these factors should also not have led to meaningful confounding. Consistent with this, we did not adjust for additional covariates and did no modeling or smoothing; causal effect estimates for the example can be calculated from just the data in Table 2 (main text).

**Table S1.**
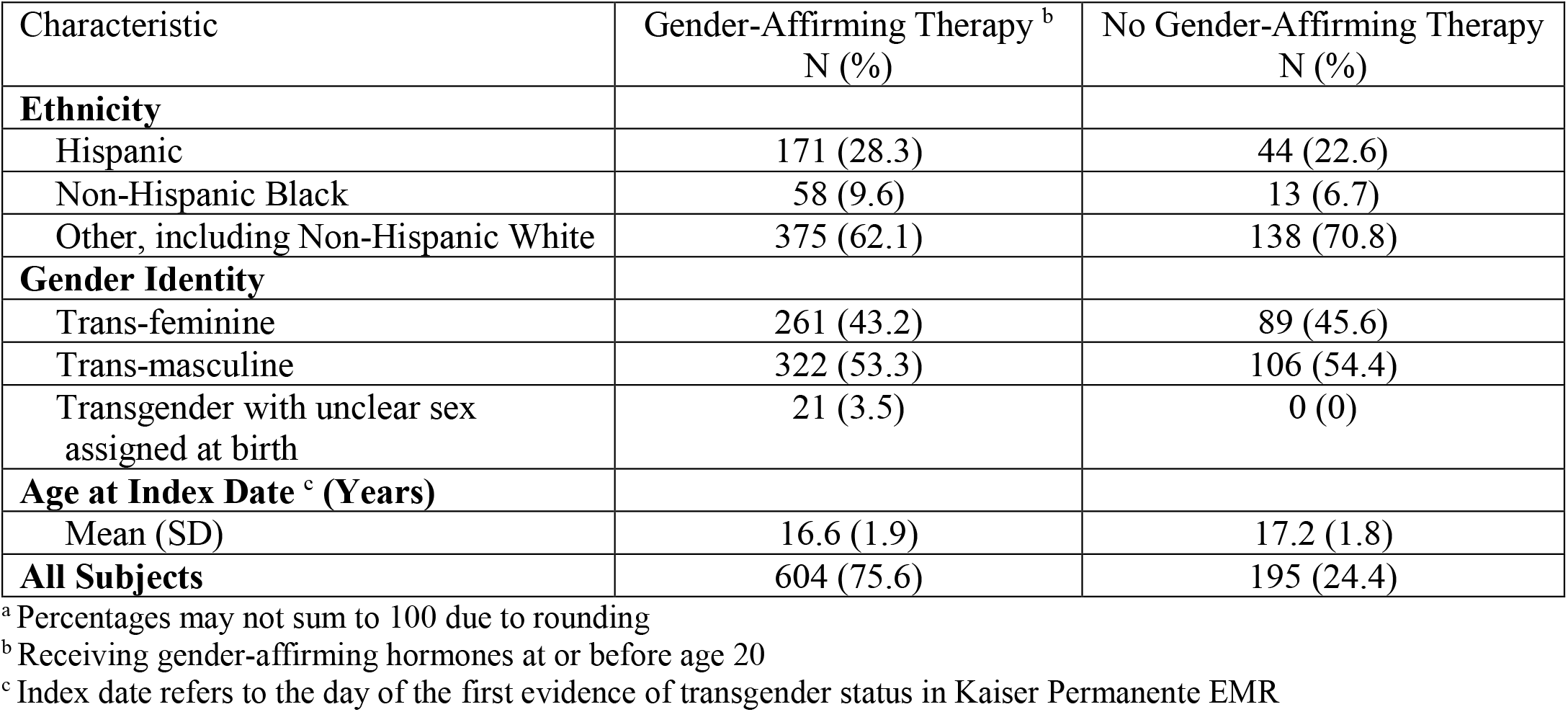
Descriptive Information for Cohort of Example in the Main Text ^a^

## Web Appendix, Figure S1

**Figure S1.**
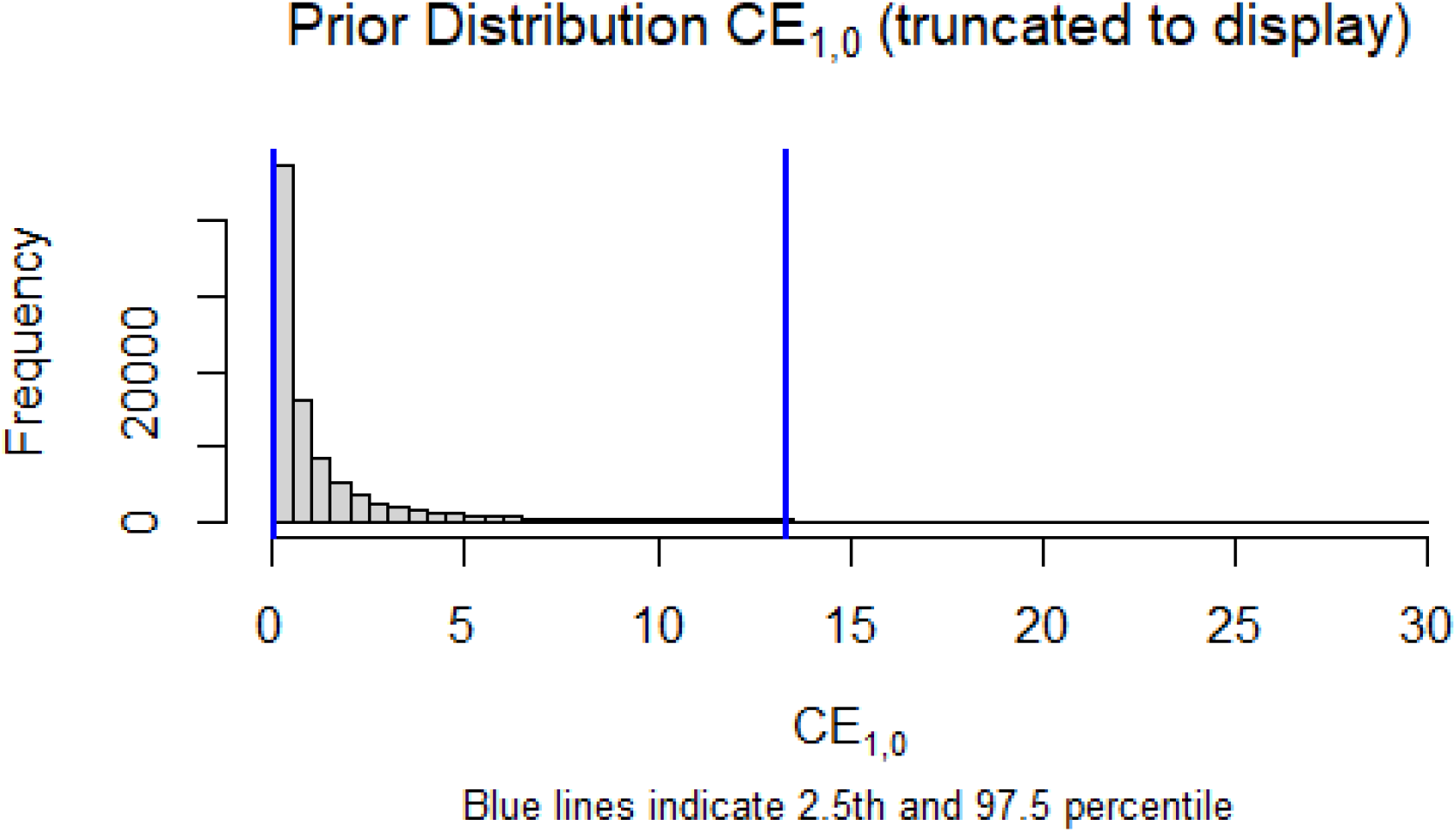
describes the prior distribution of *CE*_2,1_ for the example in the main text; see also Web Appendix S1. Values of *CE*_2,1_ were truncated at 30 to display.

## Web Appendix, Figure S2

**Figure S2.**
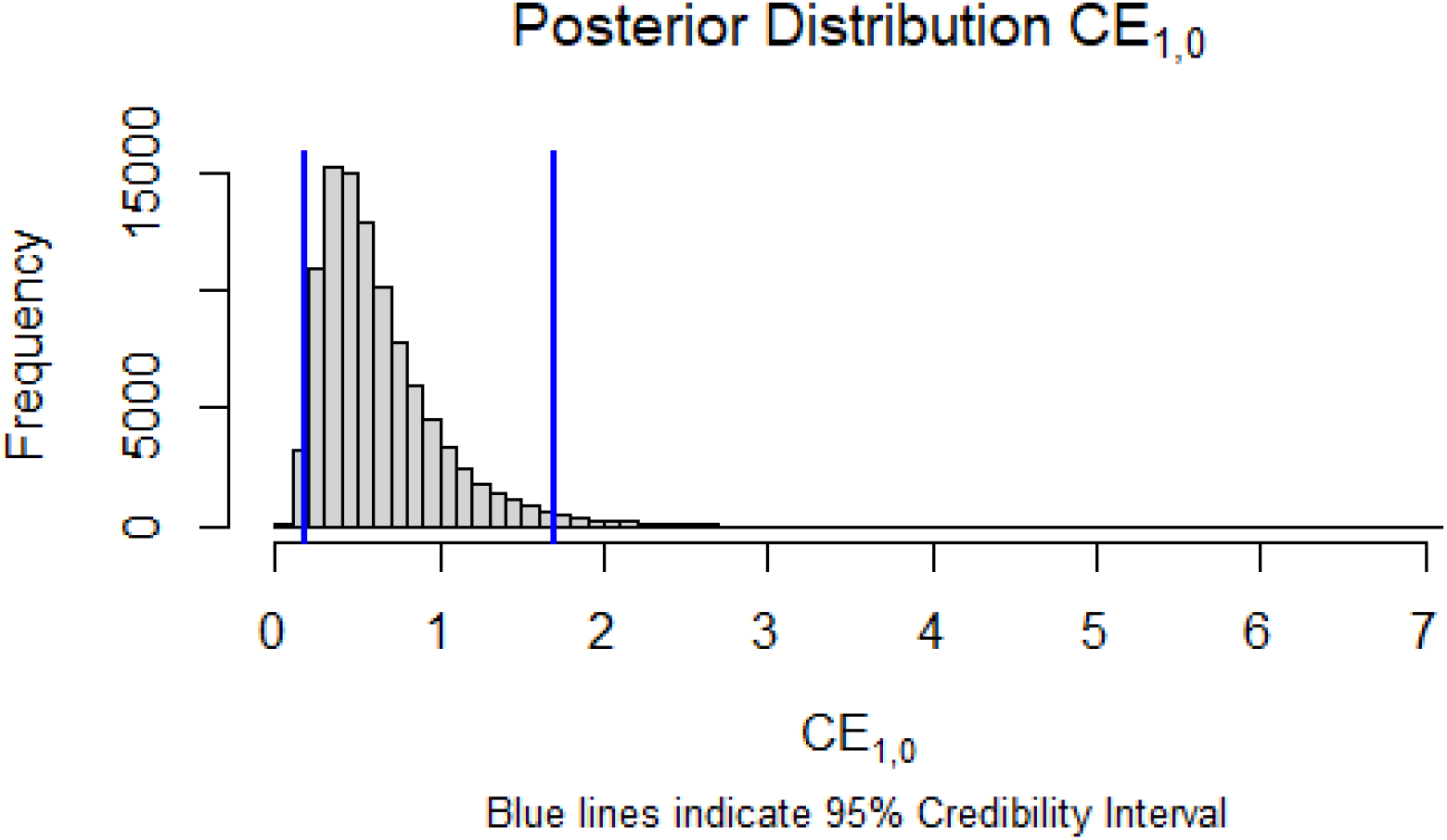
describes the posterior distribution of *CE*_2,1_ for the example in the main text; see also Web Appendix S1. (Values not truncated.)

## Web Appendix, Figure S3

**Figure – S3.**
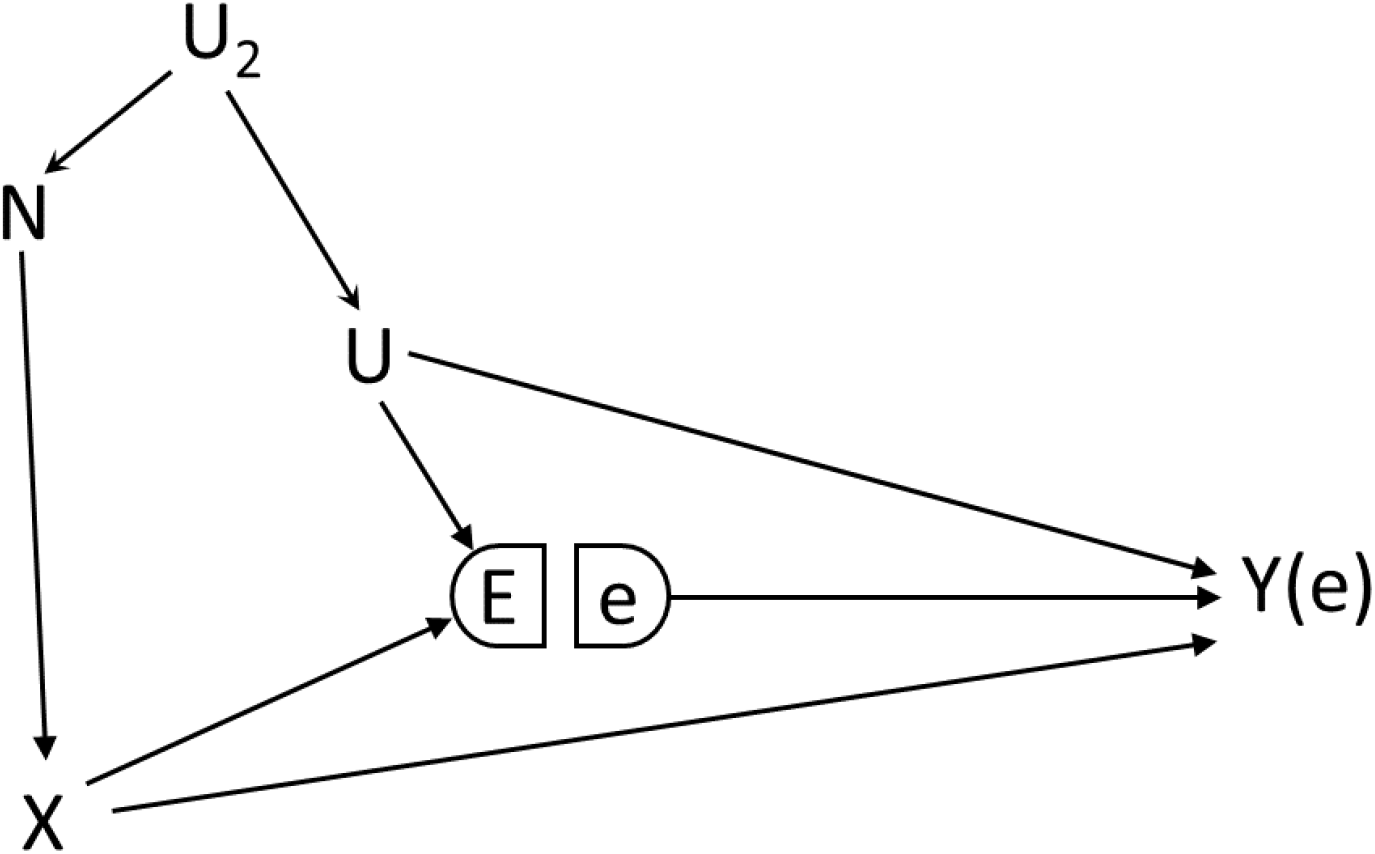
E and N have different causes in common with Y, but N can still serve as a negative control exposure; the equi-distributional assumption (A5a) or (A5b) may be less plausible.

## References

1. Lash TLV, Tyler J., Haneuse S, Rothman KJ. Modern epidemiology. 4th ed. Philadelphia: Wolters Kluwer Health/Lippincott Williams & Wilkins; 2021.

2. Shi X, Miao W, Tchetgen Tchetgen EJ. A selective review of negative control methods in epidemiology. Current Epidemiology Reports. 2020:1–13.

3. Yerushalmy J. The relationship of parents’ cigarette smoking to outcome of pregnancy— implications as to the problem of inferring causation from observed associations. International journal of epidemiology. 2014;43(5):1355–1366.

4. Taylor AE, Smith GD, Bares CB, Edwards AC, Munafò MR. Partner smoking and maternal cotinine during pregnancy: implications for negative control methods. Drug and alcohol dependence. 2014;139:159–163.

5. Flanders WD, Klein M, Darrow LA, et al. A Method to Detect Residual Confounding in Spatial and Other Observational Studies. Epidemiology (Cambridge, Mass). 2011;22(6):823.

6. Flanders WD, Klein M, Strickland M, et al. A method of identifying residual confounding and other violations of model assumptions. Epidemiology. 2009;20(6):S44–S45.

7. Jackson LA, Jackson ML, Nelson JC, Neuzil KM, Weiss NS. Evidence of bias in estimates of influenza vaccine effectiveness in seniors. International journal of epidemiology. 2006;35(2):337–344.

8. Lipsitch M, Tchetgen Tchetgen EJ, Cohen T. Negative Controls: A Tool for Detecting Confounding and Bias in Observational Studies. Epidemiology. 2010;21(3):383–388.

9. Swanson SA, Hernán MA, Miller M, Robins JM, Richardson TS. Partial identification of the average treatment effect using instrumental variables: review of methods for binary instruments, treatments, and outcomes. Journal of the American Statistical Association. 2018;113(522):933–947.

10. Flanders WD, Strickland MJ, Klein M. A new method for partial correction of residual confounding in time-series and other observational studies. American journal of epidemiology. 2017;185(10):941–949.

11. Tchetgen Tchetgen EJ. The control outcome calibration approach for causal inference with unobserved confounding. American journal of epidemiology. 2014;179(5):633–640.

12. Wang J, Zhao Q, Hastie T, Owen AB. Confounder adjustment in multiple hypothesis testing. Annals of statistics. 2017;45(5):1863.

13. Jacob L, Gagnon-Bartsch JA, Speed TP. Correcting gene expression data when neither the unwanted variation nor the factor of interest are observed. Biostatistics. 2016;17(1):16–28.

14. Miao W, Geng Z, Tchetgen Tchetgen EJ. Identifying causal effects with proxy variables of an unmeasured confounder. Biometrika. 2018;105(4):987–993.

15. Shi X, Miao W, Nelson JC, Tchetgen Tchetgen EJ. Multiply robust causal inference with double-negative control adjustment for categorical unmeasured confounding. Journal of the Royal Statistical Society: Series B (Statistical Methodology). 2020;82(2):521–540.

16. Kuroki M, Pearl J. Measurement bias and effect restoration in causal inference. Biometrika. 2014;101(2):423–437.

17. Lash TL, VanderWeele TJ, Haneuse S, Rothman KJ. Bias Analysis (Chapter 21). In: Modern Epidemiology. 4th ed. Philadelphia: Wolters Kluwer Health/Lippincott Williams & Wilkins; 2021:711–754.

18. Richardson TS, Robins JM. Single World Intervention Graphs (SWIGs): A Unication of the Counterfactual and Graphical Approaches to Causality. Working Paper Number 128, Center for Statistics and the Social Sciences, University of Washingtion. 2013:http://www.csss.washington.edu/Papers/wp128.pdf.

19. Sofer T, Richardson DB, Colicino E, Schwartz J, Tchetgen Tchetgen E. On negative outcome control of unobserved confounding as a generalization of difference-in-differences. Statistical science: a review journal of the Institute of Mathematical Statistics. 2016;31(3):348.

20. Miao W, Shi X, Tchetgen Tchetgen EJ. A confounding bridge approach for double negative control inference on causal effects. arXiv preprint arXiv:180804945. 2018.

21. Flanders WD, Khoury M. Indirect assessment of confounding: graphic description and limits on effect of adjusting for covariates. Epidemiol. 1990;1(3):239–246.

22. Yanagawa T. Case-control studies: assessing the effect of a confounding factor. Biometrika. 1984;71(1):191–194.

23. Miettinen OS. Components of the crude risk ratio. American Journal of Epidemiology. 1972;96(2):168–172.

24. Greenland S. Multiple-bias modelling for analysis of observational data. Journal of the Royal Statistical Society: Series A (Statistics in Society). 2005;168(2):267–306.

25. Mak J, Shires DA, Zhang Q, et al. Suicide attempts among a cohort of transgender and gender diverse people. American journal of preventive medicine. 2020;59(4):570–577.

26. Goodman M, Nash R. Examining health outcomes for people who are transgender. Washington, DC: Patient-Centered Outcomes Research Institute (PCORI) https://doiorg/1025302/22019 AD. 2018;12114532.

27. Flanders WD, Eldridge RC. Summary of relationships between exchangeability, biasing paths and bias. European journal of epidemiology. 2015;30(10):1089–1099.

28. Hernán MA. A definition of causal effect for epidemiology. J Epidemiol Community Health. 2004;58:265–271.

29. Greenland S, Robins J. Identifiability, exchangeability, and epidemiologic confounding. Int J Epidemiol. 1986;15:413–419.

30. Gustafson P. On model expansion, model contraction, identifiability and prior information: two illustrative scenarios involving mismeasured variables. Statistical science. 2005;20(2):111–140.

## References used in Web material

1. Lipsitch M, Tchetgen E, Cohen T. Negative Controls: A Tool for Detecting Confounding and Bias in Observational Studies. Epidemiology. 2010;21(3):383–388.

